# Time Windows Voting Classifier for COVID-19 Mortality Prediction

**DOI:** 10.1101/2021.07.02.21259934

**Authors:** Tiong-Thye Goh, MengJun Liu

## Abstract

**Background:** The ability to predict COVID-19 patients’ level of severity (death or survival) enables clinicians to prioritise treatment. Recently, using three blood biomarkers, an interpretable machine learning model was developed to predict the mortality of COVID-19 patients. The method was reported to be suffering from performance stability because the identified biomarkers are not consistent predictors over an extended duration.

**Methods:** To sustain performance, the proposed method partitioned data into three different time windows. For each window, an end-classifier, a mid-classifier and a front-classifier were designed respectively using the XGboost single tree approach. These time window classifiers were integrated into a majority vote classifier and tested with an isolated test data set.

**Results:** The voting classifier strengthens the overall performance of 90% cumulative accuracy from a 14 days window to a 21 days prediction window.

**Conclusions:** An additional 7 days of prediction window can have a considerable impact on a patient’s chance of survival. This study validated the feasibility of the time window voting classifier and further support the selection of biomarkers features set for the early prognosis of patients with a higher risk of mortality.

---

There is an urgent need to assess the likelihood of a COVID-19 patient dying from the disease and to support the decisions for prioritising treatment and resource planning in the hospital. Recently Yan et al. (2020) and his team used machine learning tools to identify three biomarkers − lactic dehydrogenase, lymphocyte and high-sensitivity C-reactive protein − reported to predict the severity of the disease in individual patients more than 10 days in advance with more than 90% accuracy. Their training set consists of 375 cases of which 201 (53.6%) survived and 174 (46.4%) died. Their algorithm was developed using the last complete blood test data. However, a recent study has suggested that the performance degraded over a longer predictive window and suffered from a high level of fluctuation (Huang et al., 2020). The assumption that the same biomarkers are consistent predictors over an extended duration is unrealistic. Biomarkers in a different time frame reveal different stages of illness as each stage is characterised by specific biochemical alterations (Marcello & Luisa, 2020). Using only the last complete blood test data suffers from information loss and lower predictive performance over an extended time window. To sustain the performance, this study proposes to use all the available data to minimise information loss by designing three classifiers based on samples over three different time windows. Subsequently, the classifiers are integrated into a majority vote classifier to optimise performance (Lam & Suen, 1997).

## MATERIALS AND METHODS

### Data Collection

For this study, we used the data in Yan et al. (2020). The medical data of all patients were collected from hospitals in Wuhan, China between 10 January and 18 February 2020 using standard case report forms that included epidemiological, demographic, clinical, laboratory and mortality outcome information (Yan et al., 2020). Only the laboratory, mortality outcome and days to outcome fields were used for modelling, validating and testing. The data consists of 375 patients with 201 (53.6%) discharged and 174 (46.4%) deceased. In total 75 blood sample features were sparingly recorded across the data set. The minimal and maximal follow-up times (from admission to hospital to death or discharge) for the patients are 0 to 35 days.

### Laboratory Methods

#### Statistical Analysis

A parametric test (t test) and nonparametric test (Mann-Whitney U test) were used for continuous variables with or without normal distribution, respectively. Missing data was handled by XGBoost during model development. A two-sided α of less than 0.05 was considered statistically significant. Statistical analyses were computed using SPSS (version 24).

#### Classifier Development

The original blood sample data set was merged into daily data for each patient. The data were partitioned into 3 sub-sample sets of 7 day windows. Data set 0 consists of the final 7 days of data with 237 samples for training and validation and 101 samples for testing. Data set 1 consists of samples from 8 to 14 days with 146 samples for training and validation and 62 samples for testing, and data set 2 consists of samples beyond 14 days with 72 samples for training and validation and 31 samples for testing. For each data set, we used the XGboost method to select the important features for the respective window and used the important features to generate a single decision tree. This process created three classifiers each representing a specific time window. Finally, we combined the three classifiers into a majority vote classifier to establish the final prediction outcome. For features extraction, XGBoost parameters setting were maximum depth equal to 4, learning rate equal to 0.2, number of tree estimators set to 150, regularisation parameter α set to 1 and ‘subsample’ and ‘colsample_bytree’ both set to 0.9. To generate the single tree, we set the maximum depth to 2 and tree estimator to 1. Figure 1 depicts the classifier development process.

**Figure 1.**
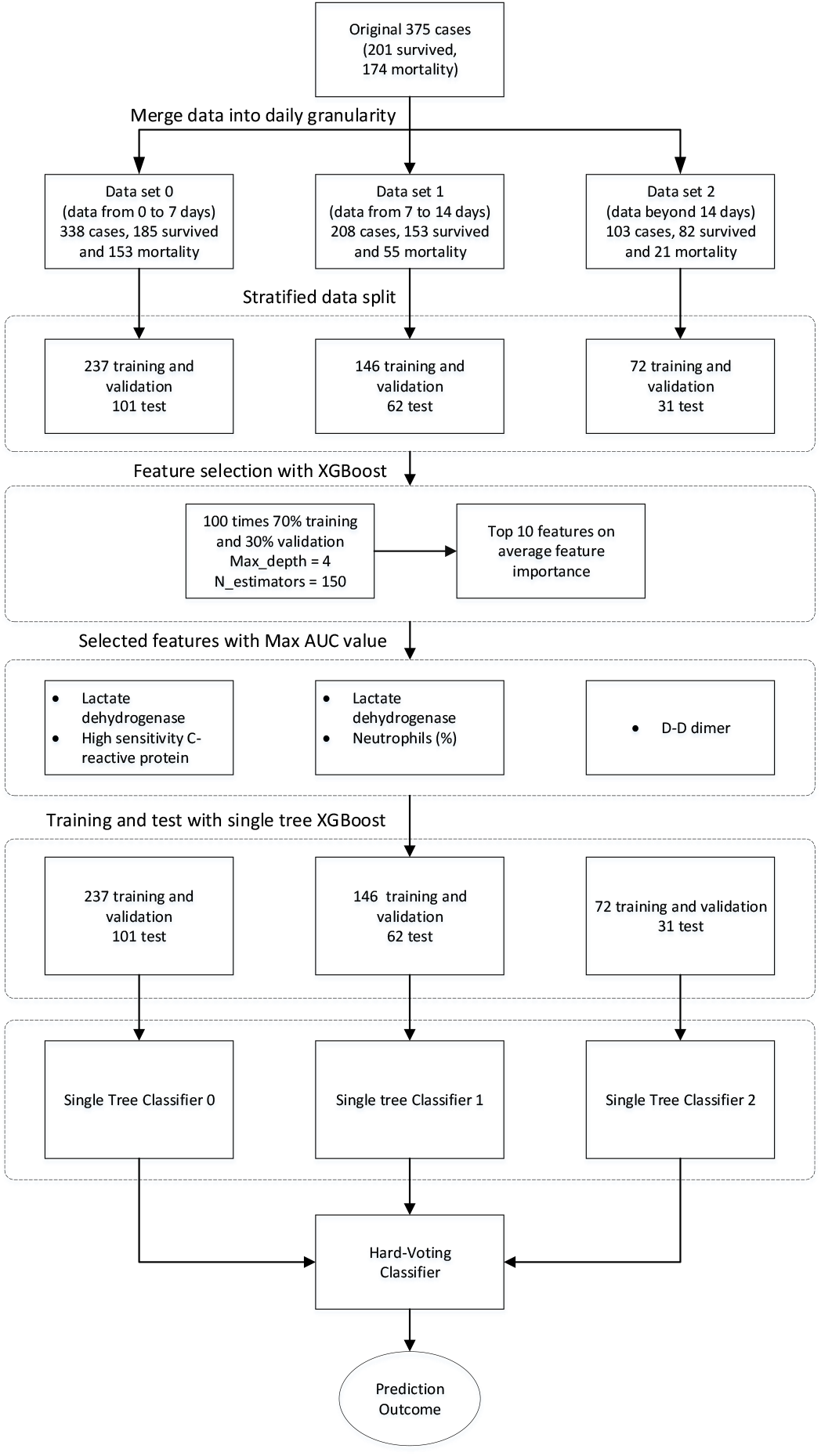
Data processing and classifier development procedures.

## Result

### Data Characteristics of COVID-19 Patients

Table 1 depicts the differences between features in the 3 data sets and patients’ outcomes. For the outcome=1 group, 13 features are significantly different (p1<0.05) between the 3 data sets, whereas the outcome=0 group 33 features are significantly different (p0<0.05) between the 3 data sets. This suggests that for the outcome=0 group, biomarkers are changing over the 3 time windows. When comparing patient outcomes between corresponding data sets, a significant majority of the features were different between the survival and the mortality group (p_dso < 0.05, p_ds1 < 0.05, p_ds2 < 0.05). These differences provide the basis for building machine learning classifiers. However, the difference statistics are not sufficient to provide information on which features are most important to provide diagnostics and predictive analysis.

**Table 1.**
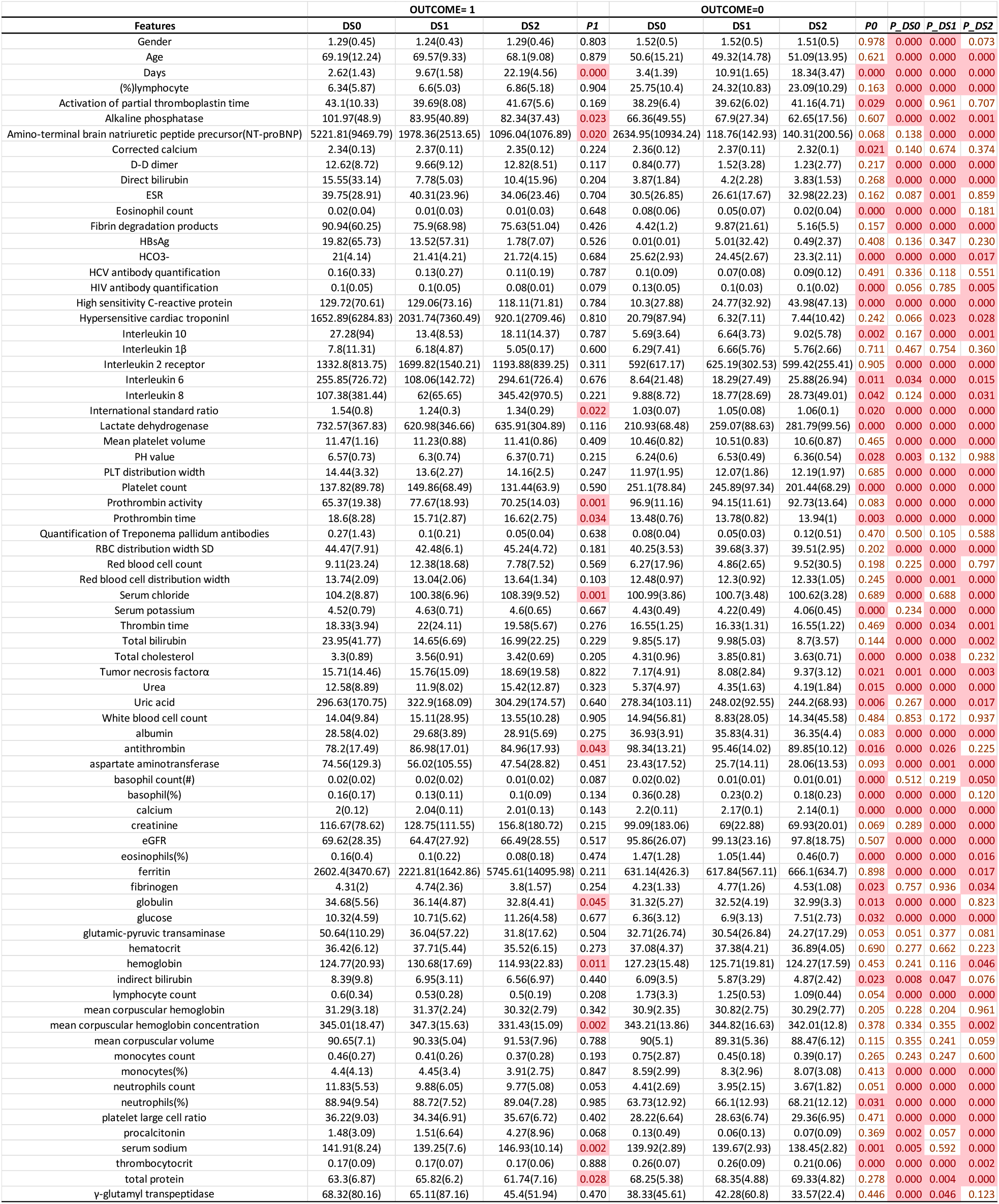
Data Sets Characteristics.

### Prognostic Factors of Severe COVID-19

The XGBoost algorithm is used to select the important features for each of the data sets. For each data set we built a single tree decision classifier for each time window. Table 2 depicts the selected prognostic features and the rules for each decision tree classifier. The biomarkers selected by the algorithm are linked to serious outcomes in patients (Ponti et al., 2020). Lactate dehydrogenase measures tissue damage associated with a wide range of disorders, including liver disease and interstitial lung disease, and C-reactive protein is a marker of inflammation or infection-reduced lung function. Lymphopenia is a common feature in patients with COVID-19 and might be a critical factor associated with disease severity and mortality (Zhao et al., 2020). Neutrophils are associated with infection, increasing the inflammation and haemorrhagic lesions in the lungs of infected patients (Hemmat et al., 2020). Neutrophils may be responsible for mortality in these severe coronavirus cases (Tomar et al., 2020). Biomolecules such as D-dimer indicate a breakdown product of blood clots. These features were identified as relevant to complications associated with COVID-19 and were dramatically increased in patients that died versus those that recovered. A study by Zhou et al. (2020) showed that D-dimer greater than 1 μg/mL could help clinicians to identify patients with a poor prognosis about 20 days in advance. D-dimer values are even higher in patients with severe COVID-19 than in those with milder forms and therefore, D-dimer measurement may be associated with evolution toward worse clinical outcomes (Lippi & Favaloro, 2020).

**Table 2.**
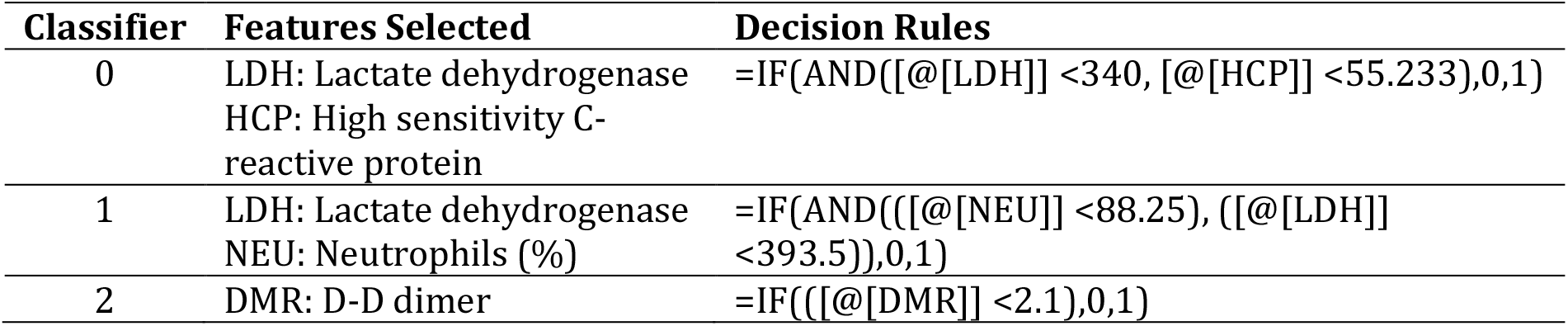
The selected prognostic features and the rules for each decision tree classifier.

### Validation of the Predictive Accuracy of the Voting Classifier

Table 3 depicts the performance of the 3 classifiers for the training, validation and the testing phases. Overall classifier 0’s average performance is better than those of classifiers 1 and 2. However when we break it down into different time windows, classifier 0’s performance degraded at time >14 days. This is the typical when using a single classifier to estimate an extended period. At time>14 days, classifier 2 performed much better than classifier 0. For all the time windows, the voting classifier was either equal or better than the best classifiers. Figure 2 shows the performance over 21 days. The voting classifier performed better than the original classifier by 5% and at above 90% accuracy.

**Table 3.**
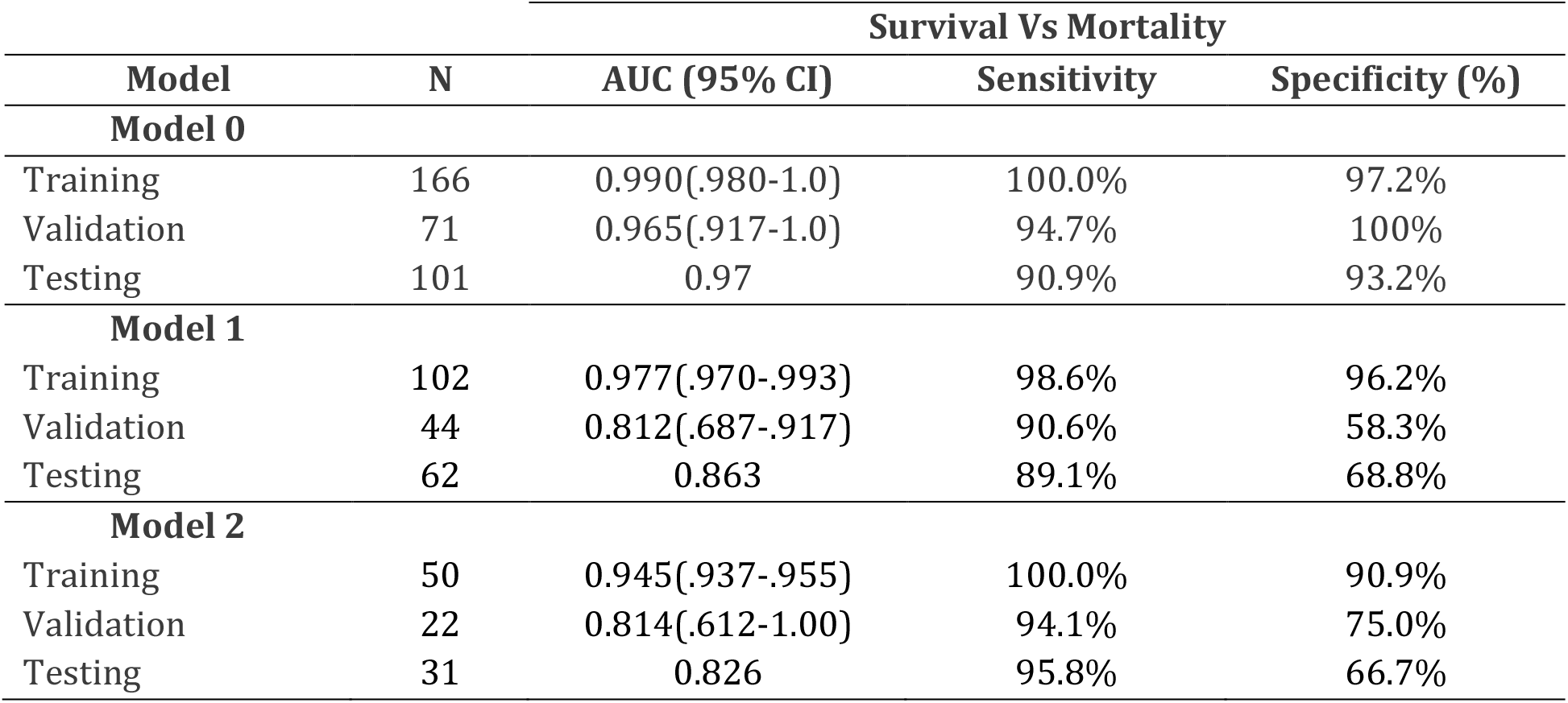
The performance of the 3 classifiers for the training, validation and the testing phases.

|  | Survival Vs Mortality |  |  |  |
| --- | --- | --- | --- | --- |
| Model | N | AUC (95% CI) | Sensitivity | Specificity (%) |
| Model 0 |  |  |  |  |
| Training | 166 | 0.990(.980-1.0) | 100.0% | 97.2% |
| Validation | 71 | 0.965(.917-1.0) | 94.7% | 100% |
| Testing | 101 | 0.97 | 90.9% | 93.2% |
| Model 1 |  |  |  |  |
| Training | 102 | 0.977(.970-.993) | 98.6% | 96.2% |
| Validation | 44 | 0.812(.687-.917) | 90.6% | 58.3% |
| Testing | 62 | 0.863 | 89.1% | 68.8% |
| Model 2 |  |  |  |  |
| Training | 50 | 0.945(.937-.955) | 100.0% | 90.9% |
| Validation | 22 | 0.814(.612-1.00) | 94.1% | 75.0% |
| Testing | 31 | 0.826 | 95.8% | 66.7% |

**Figure 2.**
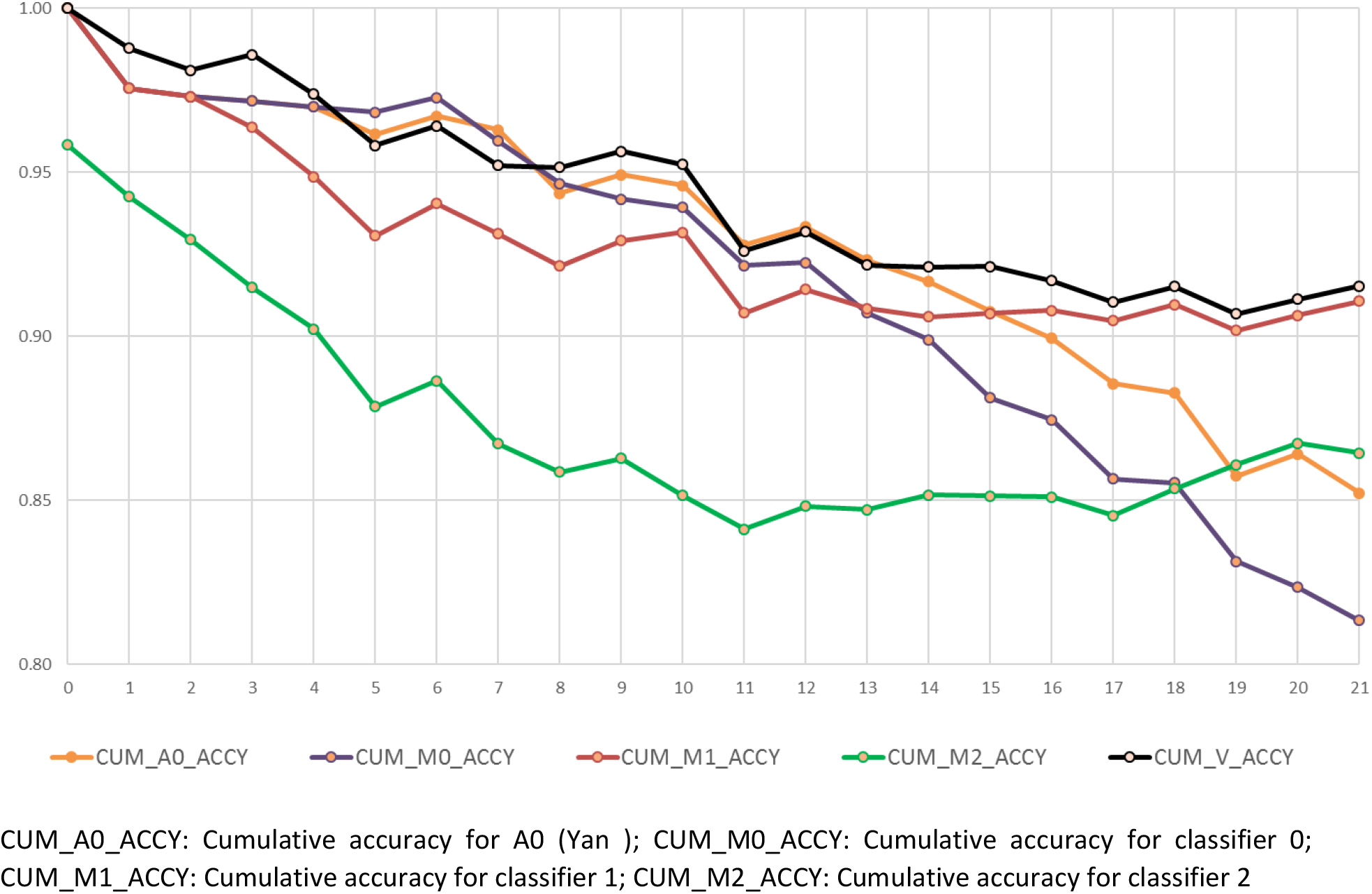
Comparing cumulative accuracy performance of classifiers over days (all available data)

## DISCUSSION

The purpose of the study is to enhance the classifier performance of Yan et al. (2020) by using all the available data instead of just the last set of complete blood data. Overall, the proposed voting classifier performs better than the original classifier from the 15 to 21 days window with 5% higher cumulative accuracy (Figure 2), 20% average accuracy (Table 4) or overall, 3.5% higher AUC (Table S).

**Table 4.** Comparison of classifiers performance based on different time window (all available data)

|  | Average Classifiers Accuracy (Mean/ SD) |  |  |  |  |
| --- | --- | --- | --- | --- | --- |
| Time Window | A0 | M0 | M1 | M2 | V |
| 0-7 days | 0.963 | 0.960 | 0.931 | 0.867 | 0.952 |
|  | 0.028 | 0.042 | 0.060 | 0.082 | 0.055 |
| 8-14 days | 0.864 | 0.830 | 0.877 | 0.834 | 0.886 |
|  | 0.108 | 0.091 | 0.128 | 0.085 | 0.131 |
| 15-21 days | 0.714 | 0.630 | 0.921 | 0.892 | 0.903 |
|  | 0.198 | 0.150 | 0.094 | 0.106 | 0.105 |

A0: Classifier based on Yan et al.; M0: Classifier 0; M1 Classifier 1; M2: Classifier 2 V: Voting Classifier

Classifier 0’s performance which is based on the ending segment of the data is compatible with the Yan et al. results as shown in Table 4 and Figure 3, especially for the 0 to 7 days window. When evaluated with a separate test data the AUCs are .888, .905,.873, and .937 for classifier 0, classifier 1, classifier 2 and the voting classifier respectively as shown in Table S. Overall the voting classifier performed as designed, especially for the 15 to 21 days window, to obtain a longer time window for the severe patients to be identified.

Figure 2 demonstrates a similar decrease in performance with distant data which is also depicted by Yan et al. (2020) in their daily performance plots. Model 1, which uses the middle segment of data, appears to be more stable than models 0 and 2. The difference in performance in each different time segment suggests that using only ending data for training may not be the best option. As can be seen from the analysis, different time segments can decrease performance due to different relevant feature indicators. When patients are admitted, the ability to establish their illness timeframe and severity improves their chance of survival.

The hard-voting classifier leverages the strength of each classifier (Lam & Suen, 1997). Figure 2 demonstrates that the overall cumulative accuracy improved from 85% (original classifier from Yan et al.) to 92% at day 21 which suggests that the simple voting classifier is a suitable approach to identify high risk patients.

Our study has several strengths. First, the decision tool is based on only 4 features, which are relatively inexpensive and easily obtained from routine blood tests. The biomarkers are linked to different stages of illness progression for poor prognosis patients. Compared to Yan et al. (2020), the proposed time window voting classifier is easy to implement and the performance is more consistent over a 21 day period. The additional test data evaluation ensures the study is rigorous.

Our study comes with certain limitations. First, the samples are retrospective and consist of 375 cases with 201 (53.6%) who survived and 174 (46.4%) who died. While the small sample size limits generalisation, it does provide a direction for further analysis when larger data sets become available. Second, a more comprehensive analysis is needed to establish the stability of the algorithm. Our method may have reduced the instability, but we do need further evidence in support. Third, the data distribution with respect to mortality would affect the performance of the algorithm which requires further validation that takes hospital context into consideration (Wang et al., 2020).

## Data Availability

data set can be obtained from
https://www.nature.com/articles/s42256-020-0180-7#Sec10

https://www.nature.com/articles/s42256-020-0180-7#Sec10

## Supplementary Data

**Notes**

## Supplement materials

**Table S.** 100 runs of single tree XGboost classifiers accuracy on test data (Mean /SD)

|  | **Classifier 0** | **Classifier 1** | **Classifier 2** | **Voting** |
| --- | --- | --- | --- | --- |
| **All Test Data** | 0.849 | 0.895 | 0.858 | 0.910 |
|  | 0.115 | 0.050 | 0.061 | 0.050 |
| **Test--DS0** | 0.969 | 0.913 | 0.852 | 0.948 |
|  | 0.016 | 0.035 | 0.040 | 0.025 |
| **Test-DS1** | 0.861 | 0.865 | 0.810 | 0.882 |
|  | 0.038 | 0.040 | 0.047 | 0.038 |
| **Test-DS2** | 0.719 | 0.907 | 0.914 | 0.900 |
|  | 0.080 | 0.058 | 0.043 | 0.056 |

## References

Hemmat, N., Derakhshani, A., Bannazadeh Baghi, H., Silvestris, N., Baradaran, B., & De Summa, S. (2020, 2020-June-09). Neutrophils, Crucial, or Harmful Immune Cells Involved in Coronavirus Infection: A Bioinformatics Study [Original Research]. Frontiers in Genetics, 11(641). https://doi.org/10.3389/fgene.2020.00641

Huang, C., Long, X., Zhan, Z., & van den Heuvel, E. (2020). Model stability of COVID-19 mortality prediction with biomarkers. medRxiv, 2020.2007.2029.20161323. https://doi.org/10.1101/2020.07.29.20161323

Lam, L., & Suen, S. (1997). Application of majority voting to pattern recognition: an analysis of its behavior and performance. IEEE Transactions on Systems, Man, and Cybernetics-Part A: Systems and Humans, 27(5), 553–568.

Lippi, G., & Favaloro, E. J. (2020, May). D-dimer is Associated with Severity of Coronavirus Disease 2019: A Pooled Analysis. Thromb Haemost, 120(5), 876–878. https://doi.org/10.1055/s-0040-1709650

Marcello, C., & Luisa, A. (2020, 24 Jun. 2020). Biochemical biomarkers alterations in Coronavirus Disease 2019 (COVID-19). Diagnosis(0), 000010151520200057. https://doi.org/10.1515/dx-2020-0057

Ponti, G., Maccaferri, M., Ruini, C., Tomasi, A., & Ozben, T. (2020). Biomarkers associated with COVID-19 disease progression. Critical reviews in clinical laboratory sciences, 1–11. https://doi.org/10.1080/10408363.2020.1770685

Tomar, B., Anders, H.-J., Desai, J., & Mulay, S. R. (2020). Neutrophils and Neutrophil Extracellular Traps Drive Necroinflammation in COVID-19. Cells, 9(6), 1383. https://doi.org/10.3390/cells9061383

Wang, C., Deng, C., & Wang, S. (2020, 2020/08/01/). Imbalance-XGBoost: leveraging weighted and focal losses for binary label-imbalanced classification with XGBoost. Pattern Recognition Letters, 136, 190–197. https://doi.org/10.1016/j.patrec.2020.05.035

Yan, L., Zhang, H.-T., Goncalves, J., Xiao, Y., Wang, M., Guo, Y., Sun, C., Tang, X., Jing, L., & Zhang, M. (2020). An interpretable mortality prediction model for COVID-19 patients. Nature Machine Intelligence, 1–6.

Zhao, Q., Meng, M., Kumar, R., Wu, Y., Huang, J., Deng, Y., Weng, Z., & Yang, L. (2020, 2020/07/01/). Lymphopenia is associated with severe coronavirus disease 2019 (COVID-19) infections: A systemic review and meta-analysis. International Journal of Infectious Diseases, 96, 131–135. https://doi.org/10.1016/j.ijid.2020.04.086

Zhou, F., Yu, T., Du, R., Fan, G., Liu, Y., Liu, Z., Xiang, J., Wang, Y., Song, B., Gu, X., Guan, L., Wei, Y., Li, H., Wu, X., Xu, J., Tu, S., Zhang, Y., Chen, H., & Cao, B. (2020, 2020/03/28/). Clinical course and risk factors for mortality of adult inpatients with COVID-19 in Wuhan, China: a retrospective cohort study. The Lancet, 395(10229), 1054–1062. https://doi.org/10.1016/S0140-6736(20)30566-3

